# Analysis of CNN features with multiple machine learning classifiers in diagnosis of monkeypox from digital skin images

**DOI:** 10.1101/2022.09.11.22278797

**Authors:** Vidit Kumar

**Affiliations:** Graphic Era Deemed to be University

## Abstract

Concerns about public health have been heightened by the rapid spread of monkeypox to more than 90 countries. To contain the spread, AI assisted diagnosis system can play an important role. In this study, different deep CNN models with multiple machine learning classifiers are investigated for monkeypox disease diagnosis using skin images. For this, bottleneck features of three CNN models i.e. AlexNet, GoogleNet and Vgg16Net are exploited with multiple machine learning classifiers such as SVM, KNN, Naïve Bayes, Decision Tree and Random Forest. Results shows that with Vgg16Net features, Naïve Bayes classifier gives highest accuracy of 91.11%.

## 1. Introduction

The fast spread of monkeypox in more than 40 nations outside of Africa has raised public health concerns. Early clinical identification of monkeypox can be difficult because of the disease’s similarities to chickenpox and measles. Even though 3–6% of people who got monkeypox during the recent outbreak died [1], isolating people who have been in contact with them and finding out who they are is important to stop the virus from spreading in the community.

Computer-aided detection of monkeypox lesions could be useful for surveillance and rapid identification of suspected cases in areas where confirmatory Polymerase Chain Reaction (PCR) tests are not easily accessible. In the case of automated disease diagnosis, deep learning algorithms have proven useful [2][3][4][5]. CNN features are also useful in other applications [6][7].

## 2. Method

In this study, we explore three deep CNN features and analyze its performance with different machine learning classifiers for monkeypox disease diagnosis. The three CNNs are AlexNet [8], GoogleNet [9] and Vgg16Net [10].

### AlexNet

It won first place at the 2012 ILSVRC. It’s an 8-layer deep network with 5 convolutional layers and 3 fully linked ones [8]. Its convolution kernel sizes are (from largest to smallest) 11×11, 5×5, 3×3, and 3×3. Its input is of 227×227×3 input. In total, there are 64,000,000 parameters that can be trained. The error rate produced by AlexNet (15.3%) was substantially greater. Additionally, it replaces the sigmoid activation function with the more effective ReLU activation function.

### Vgg16Net

Unlike AlexNet, this network employs a set of 3×3 kernels [10]. For a given receptive field range, the effect of using multiple small convolution kernels is preferred over using a single large convolution kernel. This is because the multi-layer nonlinear layer can increase the network depth, allowing for the learning of more complex patterns at a reduced computational cost.

### GoogleNet

Compared to alexnet, this CNN is more in-depth, and it adds the idea of an inception block; as a result, it placed first in the 2014 ILSVRC [9]. There are several convolutions in each Inception module, with kernel sizes of 1×1, 3×3 and 5×5 in use. Interleaving 1×1 convolutional layers accomplishes dimensionality reduction in the feature space. There are nine inception modules in total, and they all link to one another in order.

We used 5 machine learning algorithms for classification purpose. These are SVM, KNN, Decision Tree, Naïve Bayes and Random Forest.

For this, Monkeypox-Skin-Lesion-Dataset [11] is used which consists of 228 original RGB images (102 monkey pox and 126 others) and 3192 augmented images (1428 monkey pox and 1764 others). We tested on Fold 1 of the dataset.

## 3. Implementation

All the experiments are done using Pytorch with Google-Colab. The augmented images are only used during training the model. The testing is done using the original images.

To evaluate the model’s performance, four measurements are selected: accuracy, precision, recall, f1-score, which are computed using (1), (2), (3), (4) and (5) respectively.

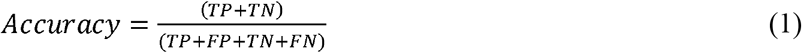

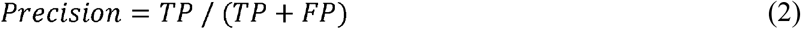

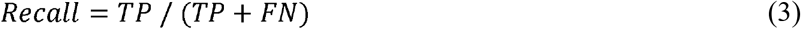

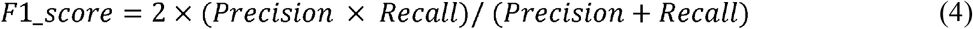

## 4. Results

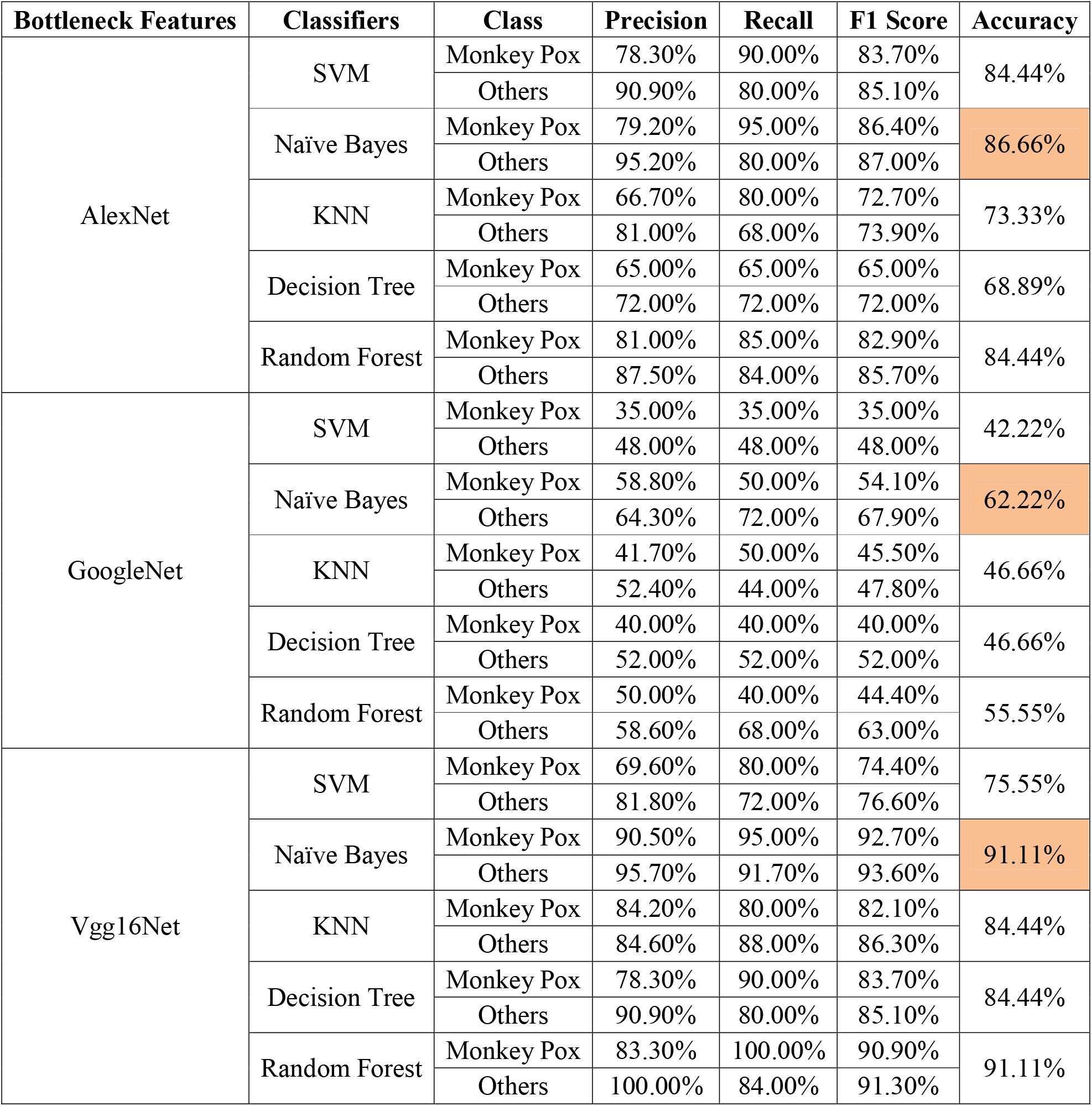

| Bottleneck Features | Classifiers | Class | Precision | Recall | F1 Score | Accuracy |
| --- | --- | --- | --- | --- | --- | --- |
| AlexNet | SVM | Monkey Pox | 78.30% | 90.00% | 83.70% | 84.44% |
|  |  | Others | 90.90% | 80.00% | 85.10% |  |
|  | Naïve Bayes | Monkey Pox | 79.20% | 95.00% | 86.40% | 86.66% |
|  |  | Others | 95.20% | 80.00% | 87.00% |  |
|  | KNN | Monkey Pox | 66.70% | 80.00% | 72.70% | 73.33% |
|  |  | Others | 81.00% | 68.00% | 73.90% |  |
|  | Decision Tree | Monkey Pox | 65.00% | 65.00% | 65.00% | 68.89% |
|  |  | Others | 72.00% | 72.00% | 72.00% |  |
|  | Random Forest | Monkey Pox | 81.00% | 85.00% | 82.90% | 84.44% |
|  |  | Others | 87.50% | 84.00% | 85.70% |  |
| GoogleNet | SVM | Monkey Pox | 35.00% | 35.00% | 35.00% | 42.22% |
|  |  | Others | 48.00% | 48.00% | 48.00% |  |
|  | Naïve Bayes | Monkey Pox | 58.80% | 50.00% | 54.10% | 62.22% |
|  |  | Others | 64.30% | 72.00% | 67.90% |  |
|  | KNN | Monkey Pox | 41.70% | 50.00% | 45.50% | 46.66% |
|  |  | Others | 52.40% | 44.00% | 47.80% |  |
|  | Decision Tree | Monkey Pox | 40.00% | 40.00% | 40.00% | 46.66% |
|  |  | Others | 52.00% | 52.00% | 52.00% |  |
|  | Random Forest | Monkey Pox | 50.00% | 40.00% | 44.40% | 55.55% |
|  |  | Others | 58.60% | 68.00% | 63.00% |  |
| Vgg16Net | SVM | Monkey Pox | 69.60% | 80.00% | 74.40% | 75.55% |
|  |  | Others | 81.80% | 72.00% | 76.60% |  |
|  | Naïve Bayes | Monkey Pox | 90.50% | 95.00% | 92.70% | 91.11% |
|  |  | Others | 95.70% | 91.70% | 93.60% |  |
|  | KNN | Monkey Pox | 84.20% | 80.00% | 82.10% | 84.44% |
|  |  | Others | 84.60% | 88.00% | 86.30% |  |
|  | Decision Tree | Monkey Pox | 78.30% | 90.00% | 83.70% | 84.44% |
|  |  | Others | 90.90% | 80.00% | 85.10% |  |
|  | Random Forest | Monkey Pox | 83.30% | 100.00% | 90.90% | 91.11% |
|  |  | Others | 100.00% | 84.00% | 91.30% |  |

## 5. Conclusion

With AlexNet features, Naïve Bayes classifier gives highest accuracy of 86.66%. With GoogleNet features, Naïve Bayes classifier gives highest accuracy of 62.22%. And, with **Vgg16Net features, Naïve Bayes classifier gives highest accuracy of 91.11%**

For future works, large scale dataset need to be explored. Also, real time efficient network to be explored for mobile based fast diagnosis. Furthermore, self-supervised approaches [12][13][14] are also needed to explore in future since labelling at large-scale can be expensive. Video based methods [15] can also be investigated for diagnosis in future.

## Data Availability

All data produced are available online at
https://github.com/mHealthBuet/Monkeypox-Skin-Lesion-Dataset

